# Limit of Detection for Rapid Antigen Testing of the SARS-CoV-2 Omicron Variant

**DOI:** 10.1101/2022.01.28.22269968

**Authors:** Sydney Stanley, Donald J. Hamel, Ian D. Wolf, Stefan Riedel, Sanjucta Dutta, Annie Cheng, James E. Kirby, Phyllis J. Kanki

## Abstract

There has been debate in the literature about the ability of antigen tests to detect the SARS-CoV-2 Omicron variant including indication on the US Food and Drug administration website that antigen tests may have lower sensitivity for the Omicron variant without provision of data or the potential scale of the issue (see https://www.fda.gov/medical-devices/coronavirus-covid-19-and-medical-devices/sars-cov-2-viral-mutations-impact-covid-19-tests-omicronvariantimpact, accessed 1/27/2022). Here we determined the limit of detection (LoD) for the Omicron variant compared with the WA1 strain used for LoD studies described in the Instructions for Use for all Emergency Use Authorization (EUA)-approved antigen tests. Using live virus (to avoid artifactual findings potentially obtained with gamma-irradiated or heat-killed virus) quantified by plaque forming units (PFU), we examined the analytical sensitivity of three antigen tests widely used in the United States: the Abbott Binax Now, the AccessBio CareStart, and LumiraDx antigen tests. We found that the 95% detection threshold (LoD) for antigen tests was at least as good for Omicron as for the WA1 strain. Furthermore, the relationship of genome copies to plaque forming units for Omicron and WA1 overlap. Therefore, the LoD equivalency also applies if the quantitative comparator is genome copies determined from live virus preparations. Taken together, our data support the continued ability of the antigen tests examined to detect the Omicron variant.

---

To bolster COVID-19 pandemic mitigation efforts, the U.S. Food and Drug Administration (FDA) issued Emergency Use Authorization (EUA) for easy-to-use rapid antigen tests instrumental for diagnosis and surveillance of SARS-CoV-2 infection (1-2). Unlike sensitive molecular tests that detect multiple SARS-CoV-2 genes, antigen tests target a singular yet genetically-conserved nucleocapsid viral protein (3-6). As the pandemic continues, some hypothesized that new SARS-CoV-2 variants might compromise antigen test performance. This concern heightened with the spread of Omicron, the B.1.1.529 variant of concern (VoC) that caused 99.5% of SARS-CoV-2 infections in the United States early 2022 (7-8). Beyond the striking 36 amino acid mutations in the spike protein, Omicron also harbors P13L, Δ31-33, R203K, and G204R nucleocapsid mutations (9). The limit of detection (LoD) of many FDA EUA antigen tests were established with gamma-irradiated or heat-inactivated preparations of the USA WA1/2020 (WA1) reference strain (13) lacking nucleocapsid mutations. This includes at-home lateral flow tests like the BinaxNOW COVID-19 Ag Card (Abbott Diagnostics Scarborough, Inc., Scarborough, ME) and the CareStart COVID-19 Antigen Home Test (Access Bio, Inc., Somerset, NJ), and the LumiraDx SARS-CoV-2 Ag Test (LumiraDx UK Ltd., Alloa, Great Britain), a microfluidic immunofluorescence assay for clinical laboratory testing (10-12). In the present study, we used cultured plaque-titered live Omicron and WA1 virus to assess differences in the LoD with the Binax, CareStart, and LumiraDx tests.

The WA1 (13) and Omicron lh01 (NCBI accession OL719310) virus were titered with standard plaque (13) and calibrated RT-qPCR (14) assays. Ten-fold serial dilutions in PBS ranging from 2.5×10^4^ to 2.5 plaque forming units (PFU)/mL were applied to swabs in 50uL volumes and tested in triplicate according to manufacturer instructions (10-12). Binax and CareStart kits contained all required consumables; iClean foam swabs (Supera CY-FS742, Houston, TX) were used with the LumiraDx test. After identifying the lowest 10-fold dilution with three replicate positive tests, we iteratively tested 3-fold dilutions around this concentration until identifying the lowest dilution (the LoD) in which at least 19 of 20 replicates (≥95%) were positive.

The LumiraDx LoD for both Omicron and WA1 was 2.5×10^2^ PFU/mL (12.5 PFU/swab or×10^6^ genome copies (gc)/swab) (Fig. 1). The Binax LoD was 8.3×10^1^ PFU/mL (4.2 PFU/swab, 3.4×10^5^ gc/swab) and 2.5×10^2^ PFU/mL (12.5 PFU/swab, 1.0×10^6^ gc/swab) for Omicron and WA1, respectively. The CareStart LoD was 2.8×10^3^ PFU/mL (1.4×10^2^ PFU/swab, 1.1×10^7^ gc/swab) and 8.3×10^3^ PFU/mL (4.2 ×10^2^ PFU/swab, 3.5×10^7^ gc/swab) for Omicron and WA1, respectively. The nearly identical relationship of PFU to genome copies for each variant indicates that the Omicron variant mutations do not change undelrying diagnostic relationships and paramaters (Figure 2). Our use of live virus, analyte volume, and swab type may explain the slight discrepancy with the manufacturers’ LoDs. Our findings are consistent with similar investigations, but these studies fell short of the FDA’s EUA requirement of 20 LoD replicates or included tests unavailable in the United States (15-17). In all, we demonstrate that the rapid antigen tests evaluated detect Omicron effectively, allaying concerns on the impact of the nucleocapsid mutations. Rapid antigen tests remain critical public health tools towards reducing SARS-CoV-2 variant transmission.

**Figure 1.**
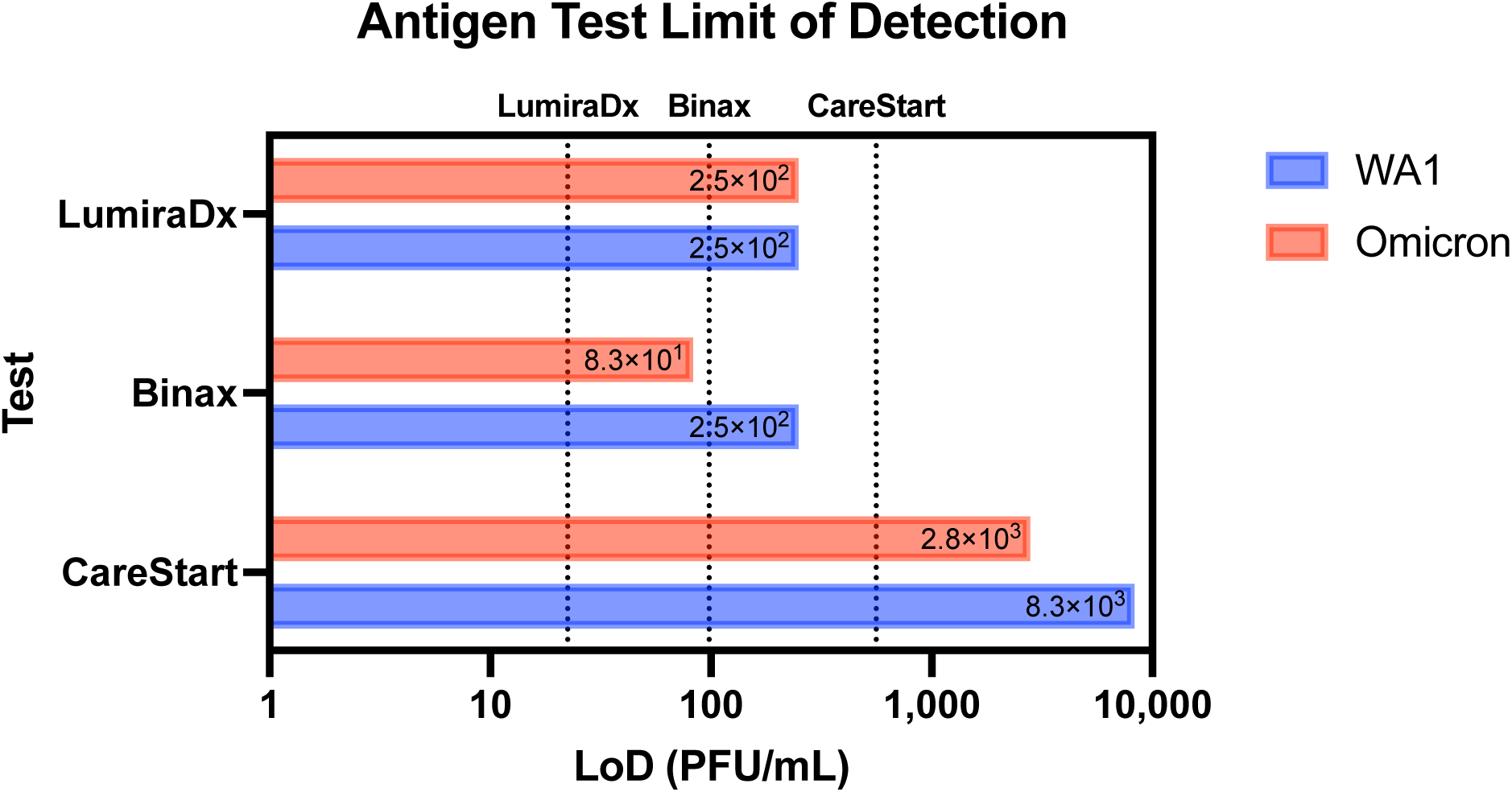
Limit of detection of the antigen tests. Limit of detection (LoD) in PFU/mL determined in our analysis (bars). Dotted lines reference the manufacturer reported LoD in respective Instructions for Use (IFU) documents (10-12), converted from TCID_50_/mL to PFU/mL by multiplying the TCID_50_/mL by 0.7, a standard conversion based on the Poisson distribution: LumiraDx (32 TCID_50_/mL, 2.2×10^1^ PFU/mL); Binax (140 TCID_50_/mL, 9.8×10^1^ PFU/mL), CareStart (800 TCID_50_/mL, 5.6×10^2^ PFU/mL).

**Figure 2.**
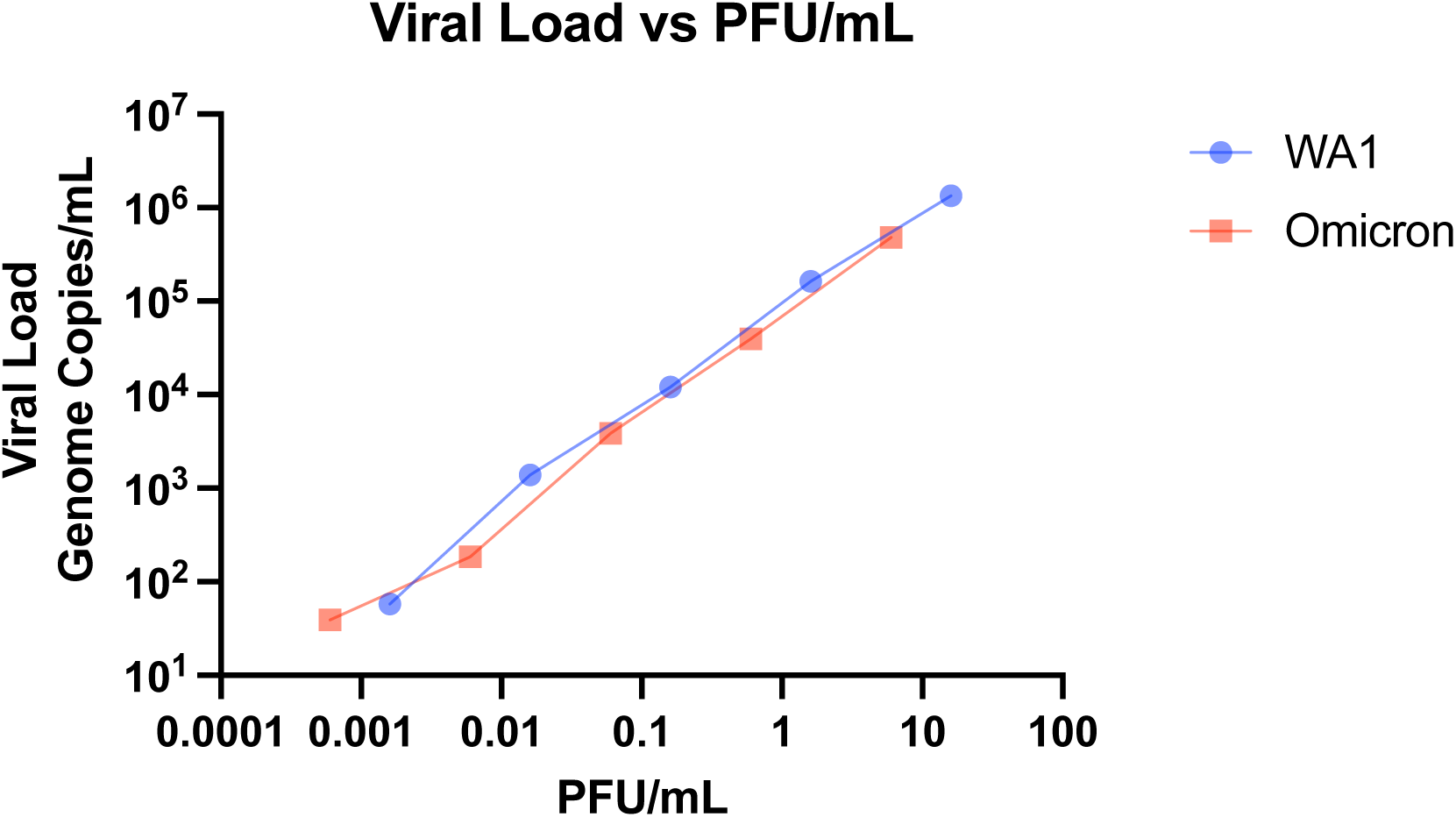
Correlation of PFU/mL and viral load in genome copies/mL. Stocks of each strain was serially diluted 10-fold in PBS and analyzed by PFU (13) and calibrated RT-qPCR assays (14). Both axes are on a Log10 scale.

## Data Availability

All data produced in the present work are contained in the manuscript.

